# Adapting COVID-19 research infrastructure to capture influenza and RSV alongside SARS-CoV-2 in UK healthcare workers winter 2022/23: Evaluation of the SIREN Winter Pressures pilot study

**DOI:** 10.1101/2024.09.08.24313279

**Authors:** Sophie Russell, Katie Munro, Sarah Foulkes, Jonathan Broad, Dominic Sparkes, Ana Atti, Jasmin Islam, Susan Hopkins, Victoria Hall, SIREN study group

## Abstract

**Background:** In 2022, with high early winter respiratory virus circulation, SIREN, a prospective healthcare worker cohort study monitoring SARS-CoV-2, ran a pilot study introducing multiplex PCR testing for SARS-CoV-2, influenza A/B, and RSV to investigate winter pressures. Three pathways were trialled: (A) on-site swabbing with local laboratory testing, (B) on-site swabbing with UKHSA-commissioned laboratory testing, and (C) postal swabbing with UKHSA-commissioned laboratory testing. Here, we compare pathways in relation to recruitment, testing coverage, participant acceptability, and UKHSA SIREN research team feedback.

**Methods:** We conducted a mixed methods evaluation using metrics of quality assurance and study fidelity (participant recruitment and retention; multiplex PCR testing timing and coverage), an adapted NIHR ‘participant in research’ feedback questionnaire, and thematic analysis of a UKHSA SIREN research team workshop.

**Results:** With 7,774 participants recruited, target recruitment (N=7,500) was achieved. Thirty-nine sites took part in the sub-study (4,289 participants). Thirty-three used pathway A, and six used pathway B. 3,485 participants enrolled to pathway C (27.8% of invitees). The median number of tests per participant was similar across pathways (6; 4; 5). However, sites using local laboratories showed a wide variation in the date they switched to multiplex testing (28^th^ November 2022 to 16^th^ March 2023). Consequently, influenza and RSV testing coverage was higher for pathways using UKHSA-commissioned laboratories (100.0% vs 45.6% at local laboratories). 1,204/7,774 (15.5%) participants completed the feedback survey. All pathways were acceptable to participants; 98.9% of postal and 97.5% of site-based participants ‘would consider taking part again’.

**Conclusion:** Transitioning SARS-CoV-2 PCR testing to include influenza and RSV was challenging to achieve rapidly across multiple sites. The postal testing pathway proved more agile, and UKHSA-commissioned laboratory testing provided more comprehensive data collection than local laboratory testing. This sub-study indicates that postal protocols are effective, adaptable at pace, and acceptable to participants.

## Introduction

In the UK, winter 2022-23 was anticipated to be particularly challenging, with relaxation of non-pharmaceutical interventions put in place to reduce COVID-19 transmission (e.g., social distancing; masking) increasing rates of circulating respiratory viruses. This effect had been seen in Australasian data where there was an increase in influenza cases, hospitalisations, and deaths (1) (2). The SARS-CoV-2 Immunity and Reinfection EvaluatioN (SIREN) study, led by the UK Health Security Agency (UKHSA), was well-placed to adapt its existing infrastructure set up during the COVID-19 pandemic to include multiplex Polymerase Chain Reaction (PCR) testing (SARS-CoV-2; influenza A, influenza B and RSV). We set up a pilot study (the SIREN Winter Pressures sub-study) to investigate the compounding burden of these three respiratory infections in healthcare workers (HCWs) (3) .

The overall design and methods of the Winter Pressures sub-study, and the wider SIREN study, have been described elsewhere (3, 4). The Winter Pressures sub-study was a nested longitudinal cohort sub-study of HCWs within the SIREN study running from 28^th^ November 2022 to 31^st^ March 2023. Expanding the study to incorporate influenza A, influenza B and RSV testing required rapid adjustment of the protocol. In the original SIREN study, participants attended hospital sites to undergo fortnightly SARS-CoV-2 PCR testing and completed an online fortnightly questionnaire on symptoms, exposures, and sick days. The Winter Pressures sub-study aimed to determine the incidence and impact of SARS-CoV-2; influenza A, influenza B and RSV on the healthcare workforce using multiplex testing. The secondary objective was to evaluate the effectiveness of seasonal influenza vaccine against infection, and power calculations required a sample size of 7,500 participants testing fortnightly to achieve this (3).

Participant recruitment proved challenging to achieve at pace, with sites facing a range of barriers including site study team capacity, laboratory availability, and access to multiplex machines. To increase the likelihood of meeting participant recruitment targets, in parallel to testing at hospital sites, a new, centrally-managed testing pathway was established for participants to undertake PCR swab via postal kits.

The sub-study comprised of three PCR testing pathways: Pathway A: fortnightly swabs were taken and tested locally at hospital sites. Pathway B: fortnightly swabs were taken locally at hospital sites but sent to UKHSA-commissioned laboratories for testing. Pathway C: fortnightly swabs were sent to participants via the post to perform at home and returned to UKHSA-commissioned laboratories for testing.

Home-based methodologies similar to Pathway C are growing in popularity for being resource-effective, reducing research costs, and reflecting a participant-centred design (5, 6). However, existing research lacks comparable information on key metrics, and does not address HCW studies where the site is also their place of work (7).

We evaluated postal and site-based testing pathways by comparing recruitment, multiplex PCR testing coverage, participant experience and acceptability, and study delivery via UKHSA SIREN research team feedback. This will inform future research protocol design, enabling comparison of postal and site-based testing options.

## Methods

We conducted a process evaluation of the SIREN Winter Pressures sub-study using mixed methods, to compare the performance and acceptability of postal and site-based testing.

We used the following quantitative and qualitative data:

1. Metrics of quality assurance and study fidelity (participant recruitment and retention, timing of introduction and coverage of multiplex PCR testing)
2. Participants research experience survey
3. UKHSA SIREN research team workshop

The methods here are informed by guidance on process evaluation from the Medical Research Council (8).

### Metrics of quality assurance and study fidelity

#### Recruitment and retention

In November 2022, all SIREN sites that intended to offer participant testing until March 2023 were invited to join the SIREN Winter Pressures sub-study via an online survey.

Sites could choose to conduct multiplex PCR swabbing and testing locally (Pathway A) or testing via UKHSA-commissioned laboratories (Pathway B). Participants at sites who joined the sub-study were sent an updated Participant Information Leaflet (PIL) informing them of the switch to multiplex testing, and were given the opportunity to withdraw from the study if they did not wish to continue. Consent was implied by continuing to undergo testing. Where possible, participants who had previously been enrolled in the study but were inactive (due to completing their period of follow-up) were re-recruited and consented to testing within the sub-study.

Participants from sites not taking part in the SIREN Winter Pressures sub-study were re-recruited and consented to join via a postal pathway (Pathway C), between 13^th^ December 2022 and 24^th^ January 2023. This pathway did not require on-site swabbing and testing.

For Pathways A and B, we recorded the number of sites joining the sub-study and the number of participants enrolled at each site when the sub-study went live.

For Pathway C, we recorded the number of participants consented and the date they consented.

To withdraw from the study, participants were required to complete a withdrawal survey. We describe recruitment, retention, and reason for withdrawal by site-based versus postal swabbing.

#### Timing of introduction and coverage of multiplex PCR testing

For sites in Pathway A, we recorded the date they switched to multiplex testing. Where this was unavailable, the date the PILs were sent to their participants was used (as a proxy for the switch to multiplex testing).

For sites in Pathway B, the date the PILs were sent to their participants was used as the date swabs should be sent to UKHSA-commissioned laboratories for multiplex testing.

For participants in Pathway C, we recorded the date the postal PCR swabs were taken. For all pathways, we calculated swab return rate as the proportion of participants who had returned at least one swab for PCR testing, and calculated the median number of tests per participant. We described the number of monoplex (SARS-CoV-2 only) PCR results and the number of multiplex (SARS-CoV-2 plus influenza and/or RSV) PCR results over time, both overall and by pathway.

### Participant survey

A bespoke adaptation of the standardised national NIHR participant research experience survey was created for participants (S1 File) (9). This was an 11-item survey that included quantitative and qualitative assessments based on participant experience. The quantitative elements included participants’ views on working with the UKHSA and site SIREN teams, the accessibility of testing, communication, and overall experience, and, for those on the postal pathway, whether they would recommend their pathway. Participants rated their views based on a 5-point Likert scale from strongly agree to strongly disagree. The survey included two free text boxes that encouraged participants to provide positive feedback or suggested improvements without limit.

Data were collated via Snapsurvey, an online survey tool, and stored securely on UKHSA internal servers. This questionnaire was sent to all participants who completed the SIREN Winter Pressures sub-study. Survey responses were anonymous. Participants were required to select their swabbing pathway within the survey to enable a comparison of participant experience between site-based and postal study designs. Site-based pathways (Pathways A and B) were grouped for this analysis as the participant experience for both pathways involved site-based swabbing. A comparison of responses was made between site-based and postal pathways using a Chi-squared test.

### UKHSA SIREN research team workshop

All researchers in the SIREN team were invited to attend a workshop to aid evaluation of the Winter Pressures sub-study. Feedback on the process of study set up and delivery was collected, and organised around core workstreams (e.g., laboratory, ethics, data collection, communications). Feedback was written up into long-form minutes that were thematically analysed.

### Thematic Analysis

All free text questions from the participant survey and SIREN research team workshop feedback minutes were thematically analysed. A pragmatic, iterative, adapted form of framework analysis was adopted (10–12).

Two independent researchers familiarised themselves with the qualitive data from both sources and independently defined key themes and sub-themes based on participant and SIREN research team responses. Themes were compared and agreed by consensus, and presented to other members of the team in the case of query or non-agreement. Themes, sub-themes and counts were summarised. The research study team then collectively discussed to finalise themes.

## Results

A summary of the key metrics from each pathway is available in Table 1.

**Table 1:**
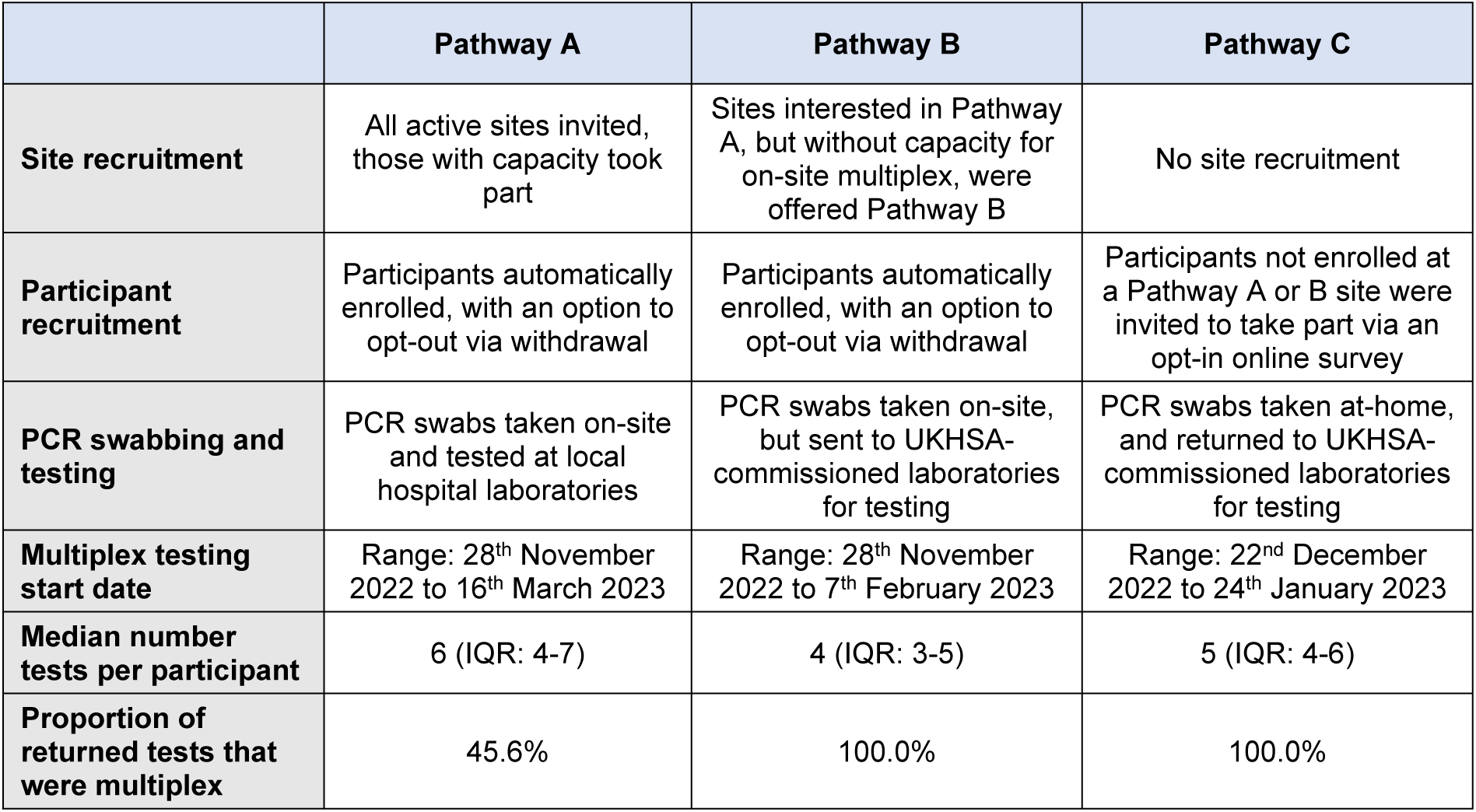
Comparison of Pathways A, B, and C.

|  | Pathway A | Pathway B | Pathway C |
| --- | --- | --- | --- |
| **Site recruitment** | All active sites invited, those with capacity took part | Sites interested in Pathway A, but without capacity for on-site multiplex, were offered Pathway B | No site recruitment |
| **Participant recruitment** | Participants automatically enrolled, with an option to opt-out via withdrawal | Participants automatically enrolled, with an option to opt-out via withdrawal | Participants not enrolled at a Pathway A or B site were invited to take part via an opt-in online survey |
| **PCR swabbing and testing** | PCR swabs taken on-site and tested at local hospital laboratories | PCR swabs taken on-site, but sent to UKHSA-commissioned laboratories for testing | PCR swabs taken at-home, and returned to UKHSA-commissioned laboratories for testing |
| **Multiplex testing start date** | Range: 28<sup>th</sup> November 2022 to 16<sup>th</sup> March 2023 | Range: 28<sup>th</sup> November 2022 to 7<sup>th</sup> February 2023 | Range: 22<sup>nd</sup> December 2022 to 24<sup>th</sup> January 2023 |
| **Median number tests per participant** | 6 (IQR: 4-7) | 4 (IQR: 3-5) | 5 (IQR: 4-6) |
| **Proportion of returned tests that were multiplex** | 45.6% | 100.0% | 100.0% |

### Recruitment and retention

Of the 65 sites invited to the sub-study, 39 (60.0%) opted to join, with 33 sites testing locally (Pathways A) and six sites testing at UKHSA-commissioned laboratories (Pathway B). In total, 4,289 participants were consented into the sub-study via sites (Pathways A: n=3,713; Pathway B: n=576).

Of the 12,549 participants invited to join the postal testing pathway (Pathway C), 3,485 (27.8%) opted to join the sub-study.

Retention across both site-based and postal pathways was high, with 4,138 (96.5%) of site-based participants, and 3,064 (87.9%) of postal participants remaining in follow-up until 31^st^ March 2023.

Overall, 572 participants withdrew from the sub-study. The most common withdrawal reason for both site-based and postal pathways was being unable to stay in the study due to workload and/or work commitments (290/572; 50.7%) (Table 2). Compared to postal participants, a higher proportion of site-based participants reported withdrawing due to logistical issues with attending appointments (14/152; 9.2% vs 5/420; 1.2%) and difficulties with accessing testing (8/152; 5.3% vs 4/420; 1.0%). Compared to site-based participants, a higher proportion of postal participants reported withdrawing due to the frequency of testing (28/420; 6.7% vs 2/152; 1.3%) and disliking the testing method (19/420; 4.5% vs 0/152; 0.0%).

**Table 2:**
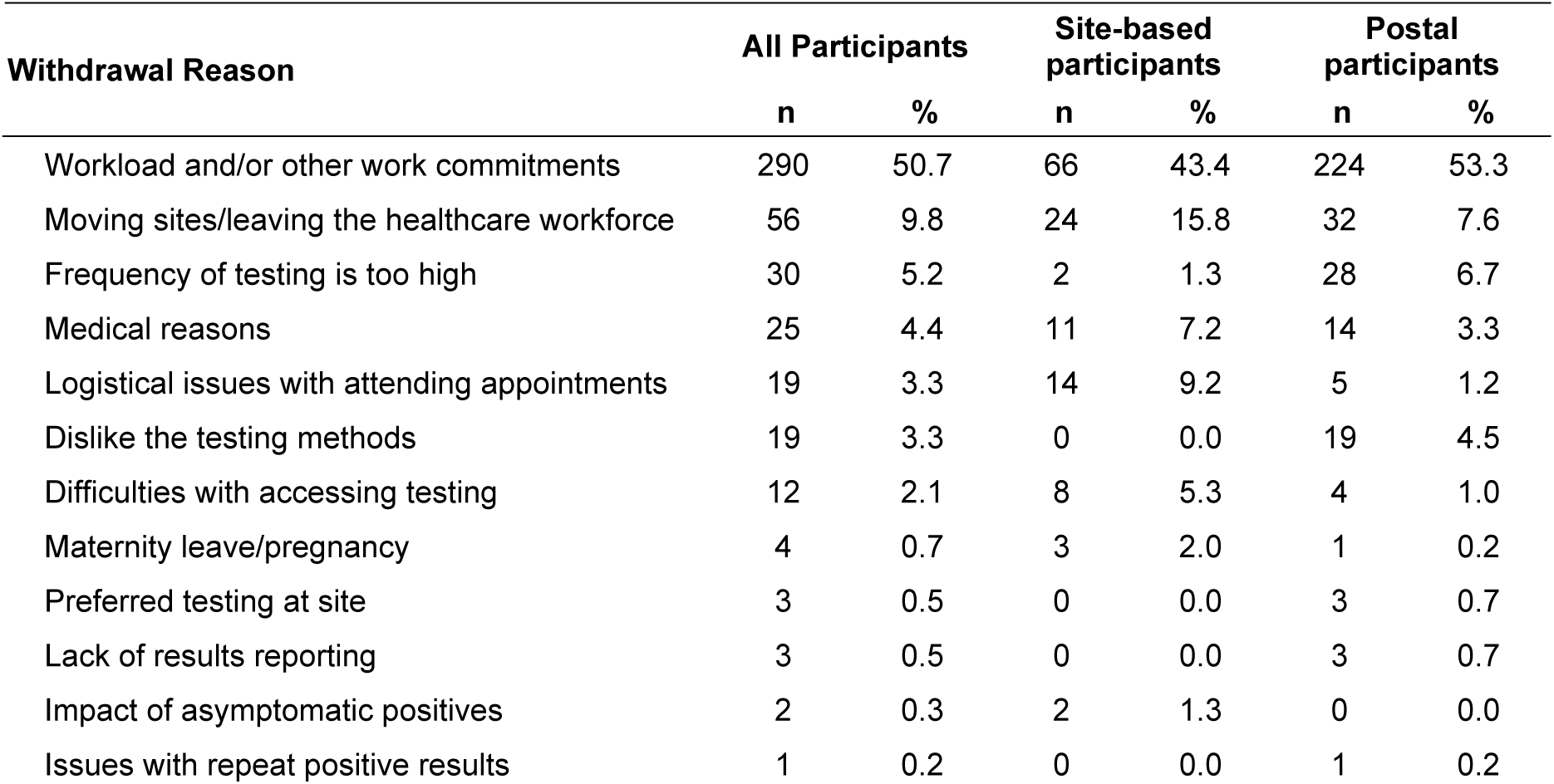
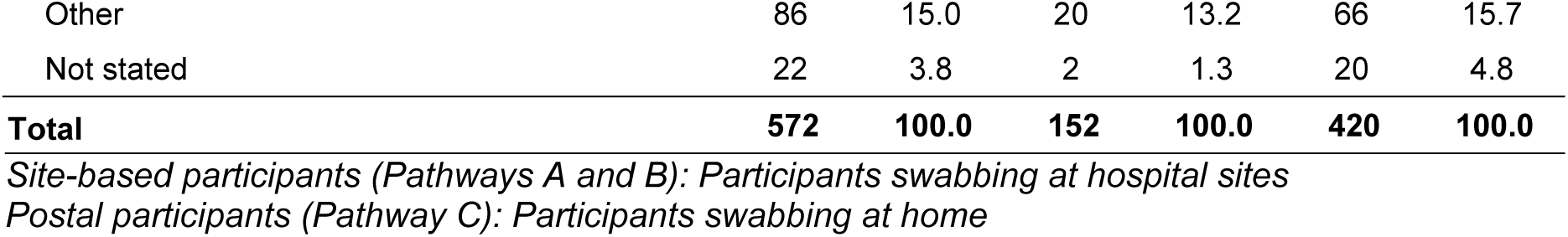
Number of participants withdrawn by withdrawal reason and pathway.

### Timing of introduction and coverage of multiplex testing

Between 28^th^ November 2022 and 31^st^ March 2023, 76.8% of participants (5,967/7,774) completed at least one SARS-CoV-2 test, and 60.9% (4,735) completed at least one multiplex test. Overall, 26,556 SARS-CoV-2 PCR tests were performed, of which 73.0% (19,396) were multiplex. Swab return rate was slightly higher for participants in Pathway C (80.5%) compared to Pathways A and B (73.7% and 73.6%, respectively). The median number of tests per participant was similar across pathways (Pathway A: 6, IQR=4-7; Pathway B: 4, IQR=3-5; Pathway C: 5, IQR=4-6). However, only 45.6% of PCR tests were multiplex in Pathway A, compared to 100.0% in Pathways B and C (Fig 1).

**Fig 1:**
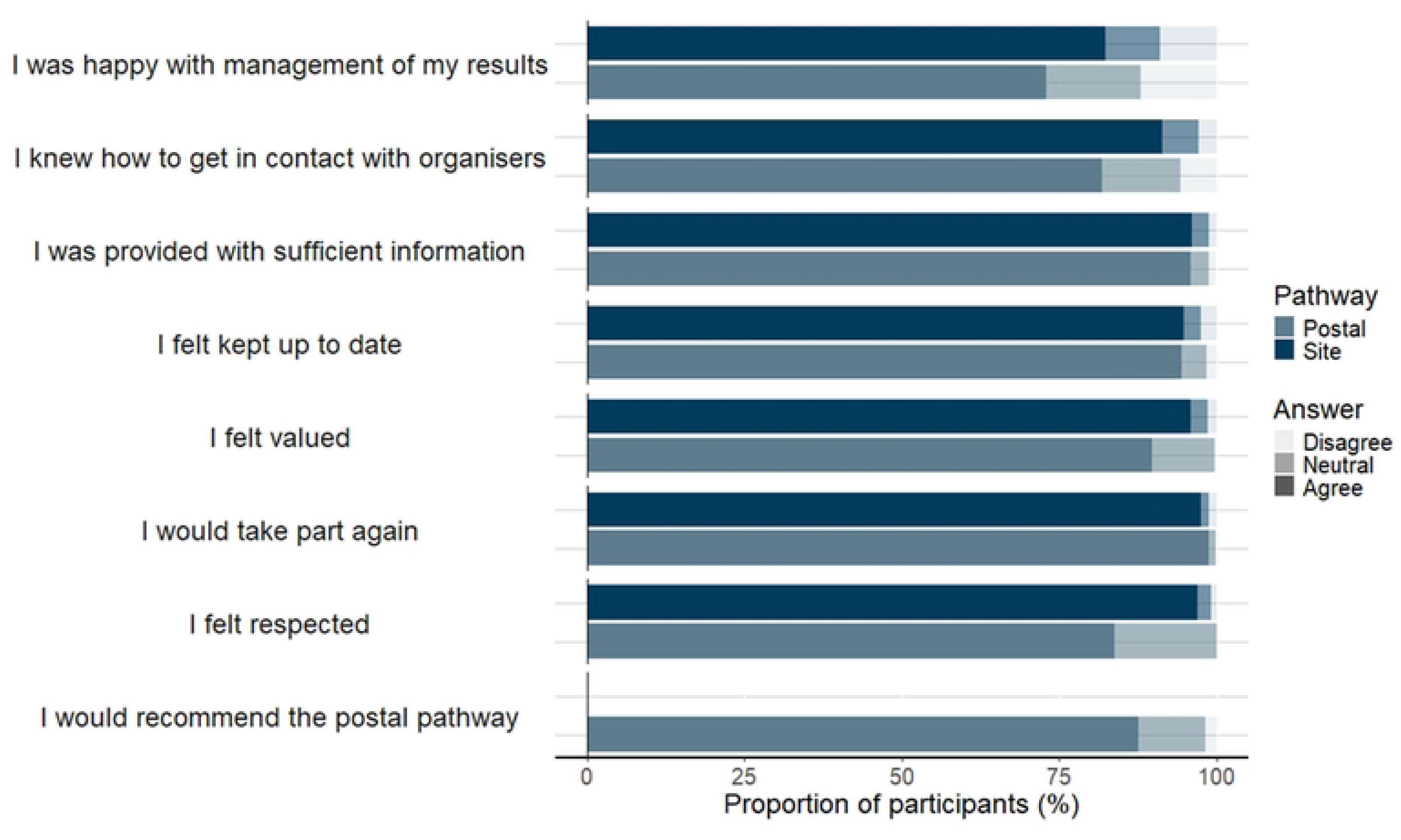
Number of participants recruited and tested per fortnight, by pathway. “Recruited” refers to the number of participants enrolled based on site/participant start dates. Pathway A: On-site PCR swabbing, swabs analysed at local hospital laboratories Pathway B: On-site PCR swabbing, swabs analysed at UKHSA-commissioned laboratories Pathway C: At-home PCR swabbing, swabs analysed at UKHSA-commissioned laboratories

On retrospective questioning of sites in Pathway A, the dates they changed from testing for SARS-CoV-2 alone to multiplex PCR ranged from 28^th^ November 2022 (which was the sub-study start date) to 16^th^ March 2023; the median date sites switched was 15^th^ December 2022. For Pathway B, the site start date ranged from 28^th^ November 2022 to 7^th^ February 2023 and all swabs sent to UKHSA-commissioned laboratories were tested on multiplex assays.

Pathway C participants were first recruited on 13^th^ December 2022, and the first samples were taken on 22^nd^ December 2022. All samples were tested on multiplex.

### Participant survey

1,204/7,774 (15.5%) participants completed the participant research experience survey about their experience of the Winter Pressures sub-study. Of these, 556 were participants at sites (Pathways A and B have been grouped as the participant experience for both pathways involved site-based swabbing), 617 were postal participants (Pathway C), and 31 did not state their pathway.

A higher proportion of site-based participants agreed to feeling valued (96.0% vs 89.8%, p<0.01), feeling respected (97.1% vs 83.9%, p<0.01), were happy with management of their results (82.3% vs 73.0%, p<0.01) and knowing who to contact (91.4% vs 81.9%, p<0.01), compared to postal participants (Fig 2). Overall, 98.2% of participants agreed they would consider taking part in research again.

**Fig 2:**
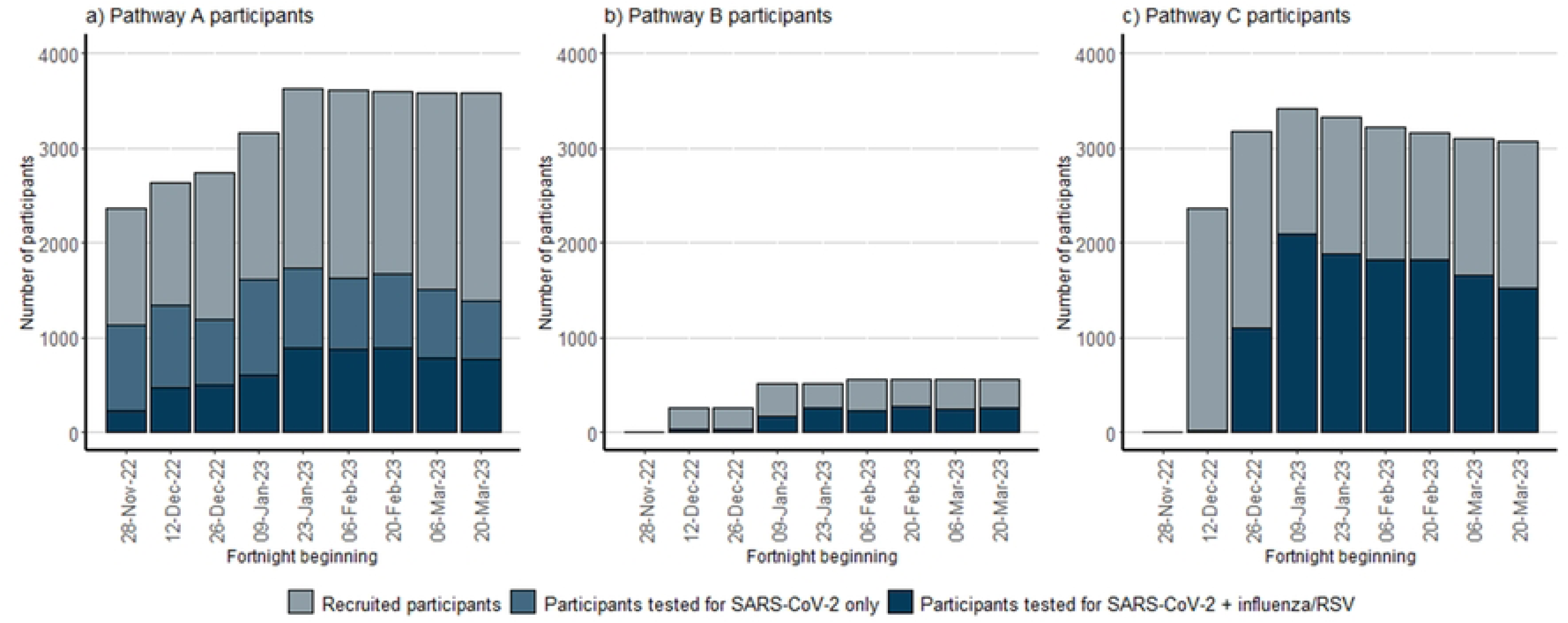
Participant research experience survey feedback from participants, divided by pathway. Site Pathway (Pathways A and B): Participants swabbing at hospital sites Postal Pathway (Pathway C): Participants swabbing at home

The three most mentioned themes in the qualitative data from the survey were ‘Taking part in SIREN creates a sense of contribution‘, ‘Logistics of the study’, and ’Participant Results‘. Within the ‘Contribution’ theme, participants felt that informing research/policy was an important factor in contributing to their participation in research. One example was:

> *“If taking part helps with the good research and you gain better understanding of these viruses, I’m happy to continue”.*

Within ‘Participant Results’, participants felt that not being informed early enough about their infection and antibody status was a disadvantage of the sub-study, compared to previous SIREN study protocols. A common suggestion was:

> *“having results more frequently”.*

Participants reported more logistical issues in the postal pathway regarding receiving and returning swabs compared to site-based swabbing. A challenge was familiarising themselves with new practises, i.e., posting swabs themselves. A suggested improvement was:

> *“having the swabs sent out earlier and in bulk as [the] first samples were hit by a postal strike”.*

Although participants did not receive incentives, this was not a deterrent to participation. For a comprehensive overview of themes and counts, see Table **3**.

**Table 3:** Participant research experience survey.

| Theme | Code | Sites | Postal | Unspecified | Total |
| --- | --- | --- | --- | --- | --- |
| Taking part in SIREN creates a sense of contribution | Belief that taking part in SIREN contributes to society and/or research and policy | 208 | 253 | 14 | 475 |
|  | Taking part in SIREN makes them feel valued | 14 | 2 | 0 | 16 |
| Participant Results | Participant knowledge of respiratory virus and antibody status | 135 | 24 | 8 | 167 |
|  | Participants received no/delayed results | 24 | 86 | 1 | 111 |
|  | Participants want detailed results/feedback | 26 | 4 | 0 | 30 |
| Logistics of the Study | Ease of study logistics for participants | 75 | 92 | 2 | 169 |
|  | Swab/survey synchronisation | 1 | 43 | 0 | 44 |
|  | Posting difficulties/convenience of sites | 0 | 29 | 0 | 29 |
|  | Received unexpected number of swabs | 1 | 25 | 0 | 26 |
|  | Issues travelling to site | 11 | 1 | 0 | 12 |
|  | Posting more convenient than on-site testing | 0 | 10 | 1 | 11 |
|  | Issues with phlebotomy | 9 | 1 | 0 | 10 |
|  | Issues with swabbing | 3 | 5 | 1 | 9 |
| Communication | Friendly/efficient site teams | 75 | 6 | 3 | 84 |
|  | Poor communication from SIREN | 22 | 52 | 0 | 74 |
|  | Good communication from SIREN | 23 | 25 | 1 | 49 |
|  | Reminders to test | 5 | 11 | 1 | 17 |
| Personal benefit | Learning about science/ results | 7 | 4 | 0 | 11 |
|  | Need for incentives | 2 | 6 | 0 | 8 |
| Surveys | Issues with survey/forms | 4 | 11 | 0 | 15 |
| Wish for SIREN to continue |  | 12 | 2 | 0 | 14 |

### UKHSA SIREN research team workshop

Twenty out of twenty-one researchers attended the workshop and provided feedback on study delivery. A variety of staff groups were represented, including laboratory, data systems, clinical, operations, and leadership. Thematic analysis of the Winter Pressures sub-study workshop identified “Communications” and “Collaboration” as the most mentioned themes (Table 4). Effective internal communications were key to sub-study delivery, including using the *“monthly meeting to update the wider team”* and the importance of gaining the *“consensus of [the] whole team”*. External communications, such *as ‘clear, considered participant communications’* and ‘*engaging the right partners at the right time’* were also important. Key challenges faced included team resourcing challenges with multiple concurrent projects leading to a ‘*need to prioritise’* and the need for clear set-up timelines, particularly when working with external partners, for example: *‘Give sites deadlines and accept that some barriers are too difficult to overcome’*.

**Table 4:** UKHSA research team workshop.

| Themes | Sub-themes | Count |
| --- | --- | --- |
| Communication | Communication between the UKHSA SIREN team and external stakeholders | 14 |
|  | UKHSA SIREN team internal communications | 11 |
| Collaboration | Engaging the most relevant collaborators | 11 |
|  | Organisational support between the SIREN team, the wider UKHSA and external partners | 9 |
|  | Clear UKHSA SIREN team roles | 4 |
| Strategy and Structures | Effective planning, process design, and contingency planning | 10 |
|  | Setting study aims and objectives | 6 |
|  | Regular agreement and discussion within the UKHSA SIREN team | 5 |
|  | Quality assurance and monitoring | 2 |
| Timescales | Includes timescales, deadlines, and parallel processes | 21 |
| Resources and Prioritisation | Includes use of wider support from UKHSA and collaborators, prioritisation, and financial processes | 15 |

## Discussion

The SIREN Winter Pressures sub-study 2022/23 involved rapid adjustment of the SIREN study protocol. Modifying the multi-centre study to switch to multiplex analysis of PCR tests proved challenging to achieve at pace, so a new postal pathway was rapidly established. This evaluation provides an opportunity to reflect and compare the site-based (A and B) and postal (C) swabbing and testing pathways.

Compared to the postal pathway, set up and delivery of site-based pathways proved time-intensive and lacked standardisation, leading to more complexity, often missing data, and creating barriers to effective timely data collection. The postal pathway enabled target recruitment to be achieved (7,774/7,500). However, with only 60.9% of participants testing for influenza, and the prolonged switch to multiplex resulting in testing starting after the influenza peak, this impacted the utility of data collected (Table 1) (13). This meant we were unable to conduct the planned analyses, including seasonal flu vaccine effectiveness. The overall return rate was similar across pathways, with a slightly higher percentage of returned kits for the postal pathway compared to site-based (80.7% vs 73.7%). This is higher than a previously reported postal PCR sample return rate of 39.9% (14).

Multiplex PCR testing coverage was highly varied for sites, with significant differences in the date when assays were transitioned from monoplex testing (Table 1). Initial recruitment figures were based on the number of consented participants that had not withdrawn at site start date. However, actual numbers of tests analysed for influenza and RSV were lower than expected for participants taking part through Pathway A due to delays in local laboratories switching to multiplex testing (Table 1). In comparison, PCR tests from Pathways B and C were sent to UKHSA-commissioned laboratories for testing, where all swabs were analysed using multiplex.

This delay in sites switching to multiplex, as in Pathway A, could be mitigated by providing longer lead-in timeframes for site study set-up and a clear deadline for switch-over. This was highlighted in the SIREN research team workshop. Feedback included the importance of engaging the right partners at the right time as crucial for set-up. However, not using sites for study delivery can reduce the time needed for study set-up, as seen in Pathway C. A shorter timeframe can reduce administrative burden, enable standardisation of testing start-date, and facilitate testing start date coinciding with the predicted peak of circulating infection levels.

A comparison of site-based (A and B) versus postal (C) pathways shows that both options were acceptable to participants. This was reflected across participant feedback, swab return rates, and retention. Participants reported positive feedback for both site-based and postal approaches. A higher proportion of participants in site-based pathways agreed to feeling valued, feeling respected, were happy with management of their results and knowing who to contact, compared to the postal pathway. Responses to open questions suggest this may be due to participants valuing friendly and efficient site teams, although both arms reported a sense of contribution (see Table **3**). Overall, 98.2% of participants agreed they would consider taking part in research again, a higher percentage than the 92% reported by the NIHR Participants in Research Survey 2022/23 (9).

Retention was high in both site-based and postal participants, at 96.5% and 87.9% respectively, which is above the 73.9% overall mean retention reported in metanalysis of longitudinal cohort studies (15). Participants undergoing postal testing were more likely to withdraw than those at sites, primarily due to more postal participants reporting that workload and other commitments prevented them from completing the study. Compared to participants on the postal pathway, site-based participants were more likely to withdraw due to logistical challenges. However, all participants took on average the same number of tests, suggesting this was not a major consideration for most.

A strength of this mixed methods evaluation of the sub-study is that it incorporates multiple sources of quantitative and qualitative data. A key limitation is the low participant survey response rate (15.5%), so feedback may reflect the views the views of the most engaged participants but be unrepresentative of the overall cohort. Additionally, while the SIREN research team feedback had input from all but one team member, it was collected in a team-led workshop rather than a formalised focus group moderated by independent researchers. However, UKHSA SIREN team debriefs have now been adapted into the team culture to support continuous improvement of study delivery.

## Conclusion

The Winter Pressures sub-study was a pragmatic adaptation of the existing SIREN protocol, which faced real-world challenges impacting data collection. While the target recruitment of participants was met, the number of returned multiplex test results did not achieve the intended large-scale seasonal coverage. This evaluation compares the three pathways used in the sub-study, and supports the postal pathway as an effective format for data collection that is acceptable to participants and the research team. The postal pathway offers greater control over the end-to-end processes, providing an adaptable and responsive format with implications for use in pandemic preparedness and for emerging public health threats.

## Data Availability

Anonymised data will be made available for secondary analysis to trusted researchers upon reasonable request.

## Acknowledgements

Our thanks go to the participants in the SIREN study and in particular those who took part in the Winter Pressures sub-study 2022/23, and completed the evaluation questionnaire. We would also like to thank the research team and site staff.

## Supporting information

**S1 File: Participant research experience survey questions**

**S2 File: SIREN study group members**

## References

1. Office UGC. COVID-19 Response: Living with COVID-19 https://www.gov.uk/government/publications/covid-19-response-living-with-covid-19/covid-19-response-living-with-covid-192022 [updated 6th May 2022].

2. Government A. Australian Influenza Surveillance Report. In: Care DoHaA, editor. Reporting fortnight: 26 September to 09 October 2022. https://www.health.gov.au/sites/default/files/documents/2022/10/aisr-fortnightly-report-no-14-26-september-to-9-october-2022.pdf2022.

3. Broad J, Sparkes D, Platt N, Howells A, Foulkes S, Khawam J, et al. Adapting COVID-19 research infrastructure to capture Influenza and Respiratory Syncytial Virus alongside SARS-CoV-2 in UK healthcare workers Winter 2022/23 and beyond: protocol for a pragmatic sub-study. medRxiv. 2023:2023.09.19.23295789.

4. Hall VJ, Foulkes S, Charlett A, Atti A, Monk EJM, Simmons R, et al. SARS-CoV-2 infection rates of antibody-positive compared with antibody-negative health-care workers in England: a large, multicentre, prospective cohort study (SIREN). The Lancet. 2021;397(10283):1459–69.

5. National Academies of Sciences E, Medicine, Health, Medicine D, Board on Health Sciences P, Forum on Drug Discovery D, et al. The National Academies Collection: Reports funded by National Institutes of Health. In: Shore C, Khandekar E, Alper J, editors. Virtual Clinical Trials: Challenges and Opportunities: Proceedings of a Workshop. Washington (DC): National Academies Press (US) Copyright 2019 by the National Academy of Sciences. All rights reserved.; 2019.

6. Ali Z, Zibert John R, Thomsen Simon F. Virtual Clinical Trials: Perspectives in Dermatology. Dermatology. 2020;236(4):375–82.

7. Rogers A DPG, Subbarayan S, et al. A systematic review of methods used to conduct decentralised clinical trials. British Journal of Clinical Pharmacology. 2022:88(6):2843–62.

8. Moore GF, Audrey S, Barker M, Bond L, Bonell C, Hardeman W, et al. Process evaluation of complex interventions: Medical Research Council guidance. bmj. 2015;350.

9. 2022/23 PRES executive summary NIHR: NIHR; 2023 [updated 19/09/2023. Available from: https://www.nihr.ac.uk/documents/202223-pres-executive-summary/34466.

10. Ritchie J, Spencer L. Qualitative data analysis for applied policy research. Analyzing qualitative data: Routledge; 2002. p. 187–208.

11. Thomas J, Harden A. Methods for the thematic synthesis of qualitative research in systematic reviews. BMC Medical Research Methodology. 2008;8(1):45.

12. Braun V, Clarke V. Using thematic analysis in psychology. Qualitative Research in Psychology. 2006;3(2):77–101.

13. UKHSA. Surveillance of influenza and other seasonal respiratory viruses in the UK, winter 2022 to 2023 2024 [Available from: https://www.gov.uk/government/statistics/annual-flu-reports/surveillance-of-influenza-and-other-seasonal-respiratory-viruses-in-the-uk-winter-2022-to-2023#:~:text=High%20levels%20of%20influenza%20activity,the%20remainder%20of%20the%20season.

14. Marchant E, Ready D, Wimbury G, Smithson R, Charlett A, Oliver I. Determining the acceptability of testing contacts of confirmed COVID-19 cases to improve secondary case ascertainment. Journal of Public Health. 2021;43(3):e446–e52.

15. Teague S, Youssef GJ, Macdonald JA, Sciberras E, Shatte A, Fuller-Tyszkiewicz M, et al. Retention strategies in longitudinal cohort studies: a systematic review and meta-analysis. BMC Medical Research Methodology. 2018;18(1):151.

